# Causal effect of loss to follow-up on mortality in a population-based tuberculosis cohort in Brazil

**DOI:** 10.64898/2026.06.29.26356808

**Authors:** Evelyn Lepka de Lima, Ana Angélica Lindoso, Giovanna Orlandi, Suely Fukasava, Catia Martinez, Julio Croda, C. Robert Horsburgh, Otavio Ranzani, Meredith B. Brooks, Jason R. Andrews

## Abstract

**Background:** Loss to follow-up (LTFU) during tuberculosis treatment is a major programmatic gap, but how much excess mortality it causes has been difficult to estimate, because individuals who disengage differ from those who do not, and they must survive long enough to be recorded as having disengaged. Quantifying this excess and identifying which populations are at greatest risk are critical to informing program design and resource allocation.

**Methods and Findings:** We followed 171,048 people aged 15 years or older starting a first course of tuberculosis treatment in São Paulo State, Brazil, between 2013 and 2023, linking surveillance and mortality registries; 20,830 (12.2%) were LTFU. Among those LTFU, 65% had received directly observed therapy (DOT). Using a clone-censor-weight target trial emulation, we contrasted two strategies over 60 months from the first day of treatment: disengagement at any point during the first six months and remaining in care throughout. A rolling landmark analysis addressed the timing of disengagement and cause-specific mortality excess. LTFU increased standardized five-year mortality by 2.23 percentage points (95% CI 1.88–2.51), with tuberculosis accounting for 61% of the excess. Relative effects were largest in younger, stably housed individuals, whose baseline mortality was low; the largest absolute excess fell on people living with HIV (4.49 percentage points, 95% CI 2.97-5.74). The excess was similar for disengagement at any point in the first four months of treatment and attenuated thereafter. Limitations include the absence from the registry of severity markers such as radiographic extent and bacillary load, so residual confounding cannot be excluded.

**Conclusions:** Despite high DOT coverage, disengagement during the first six months of tuberculosis treatment was common and increased five-year mortality under the stated identification assumptions, with the excess concentrated in deaths attributed to tuberculosis. Interventions that provide support throughout treatment should be tested for their effectiveness in sustaining engagement and reducing mortality.

## Introduction

Tuberculosis remains a major cause of infectious disease mortality globally, accounting for an estimated 1.23 million deaths in 2024. (1) Achieving progress in reducing this burden depends critically on maintaining high rates of completion of curative therapy. Despite the availability of effective drug regimens for decades, many high-burden countries have struggled to translate the high cure rates observed in clinical trials and controlled settings into everyday practice. (2) Mortality trends in the WHO Region of the Americas further highlight this challenge, with limited progress overall and increasing mortality in several countries, including Brazil, since 2015. (1)

Loss to follow-up (LTFU) over the standard six months of tuberculosis treatment represents an important programmatic gap in tuberculosis care and a major contributor to adverse outcomes. When LTFU occurs, consequences include *Mycobacterium tuberculosis* drug resistance, ongoing transmission, and increased mortality (1). However, quantifying these effects presents several challenges. LTFU is not a random event, and a range of demographic and clinical characteristics have been associated with increased risk, including male sex, younger age, lower educational attainment, HIV co-infection, prior tuberculosis, and substance use. (1, 2)

Estimating the impact of LTFU on mortality is therefore complicated by confounding, as factors influencing disengagement from care also affect subsequent outcomes. (3) These effects can act in multiple directions: individuals with greater social vulnerabilities may be more likely to interrupt treatment and have poor outcomes, while individuals with more severe disease, who are at greatest risk of death, might be less likely to discontinue therapy than those with less advanced disease and few symptoms.

Comparisons between individuals who interrupt treatment and those who complete it are also subject to several distinct forms of immortal-time bias (4). When follow-up is counted from treatment initiation, an individual can be classified as LTFU only by surviving long enough to disengage from care, whereas those who die while still in treatment are recorded as on-treatment deaths and can never enter the LTFU group. The LTFU group is therefore guaranteed to have survived up to the moment of disengagement and crediting that survival time to it makes LTFU appear spuriously protective over short follow-up times (5). Further, because programmatic LTFU is declared only after a specified period of absence, individuals classified as LTFU must survive that additional interval, a requirement that the on-treatment comparator does not have. Conversely, defining the comparator as individuals who complete therapy introduces immortal-time bias from the other direction, since treatment completion is itself a post-baseline outcome attainable only by surviving to the end of treatment.

Prior studies have consistently found higher mortality after LTFU (6–11), but most compared individuals lost to follow-up with those recorded as cured or as having completed treatment (6, 7, 9, 11), a group that cannot contain anyone who died on therapy. None has addressed these biases systematically, so the magnitude of the excess mortality caused by disengagement remains unknown. Quantifying the effects of LTFU on mortality and identifying the groups at greatest risk of harm from LTFU are both important for prioritizing resources and developing effective strategies to address this problem.

We therefore aimed to address these methodological challenges and estimate the effect of LTFU on mortality, under explicitly stated identification assumptions, using a large population-based cohort linking tuberculosis surveillance and mortality data over long-term follow-up. We set out to answer three questions: how much excess mortality is attributable to disengagement; whether that effect depends on when during therapy disengagement occurs; and in which populations the absolute excess is largest. We additionally examined the causes of death underlying the excess to assess whether it is consistent with a causal pathway through interrupted therapy.

## Methods

### Study design and data sources

We conducted a retrospective cohort study using data from TBweb, the electronic tuberculosis surveillance system of the São Paulo State Tuberculosis Program, and the Brazilian Mortality Information System (SIM). TBweb is an electronic surveillance platform used to notify and monitor all individuals diagnosed with tuberculosis in São Paulo State. The system is integrated with SINAN, the national notifiable diseases database, and uses the SINAN identification number as a unique person-level identifier, enabling longitudinal follow-up and linkage of multiple tuberculosis episodes for the same individual. SIM is Brazil’s national mortality registry, which records all deaths nationwide. Death certificates follow the international format, and the underlying cause of death is assigned by trained coders based on the reported sequence of conditions. The quality of SIM has recently been evaluated: registration completeness has improved over time and the proportion of deaths assigned an ill-defined or otherwise uninformative underlying cause fell substantially between 2000 and 2015, with the remaining shortfalls concentrated in the North and Northeast rather than in São Paulo State (12, 13). The study extract included the underlying cause of death; the contributing conditions recorded on the remaining lines of the certificate were available from the same linkage and are used in a secondary analysis. TBweb and SIM records were probabilistically linked to identify deaths during follow-up (Appendix §2). The study included all tuberculosis notifications recorded in TBweb between January 1, 2013, and December 31, 2023, with administrative censoring on December 31, 2024 to allow at least 1-year of follow-up. Data extraction and linkage were performed in January 2025.

### Study population

The study cohort included individuals aged 15 years or older at the time of their first new tuberculosis notification in São Paulo State, Brazil, between 2013 and 2023. Each individual contributed a single episode: their earliest new-case notification, excluding notifications later reclassified as not tuberculosis or lacking a recorded treatment outcome. Whether that episode ended in LTFU defined the exposure. We excluded individuals with missing or implausible dates (e.g. death occurring before treatment initiation date), administrative transfers outside São Paulo State or Brazil, or no recorded treatment start date. Later notifications did not contribute exposure, but were used to ascertain cause of death. All counts refer to individuals, not notifications.

### Exposures and outcomes

LTFU during the first tuberculosis episode was the exposure of interest. Under Brazilian national guidelines, LTFU is defined as treatment interruption for at least 30 consecutive days (14). Therefore, the date recorded in the registry is the LTFU declaration date: the first day on which LTFU can be classified, after the individual has gone at least 30 days without documented medication use. To approximate the date on which treatment was actually interrupted, we defined the interruption date as 30 days before the declaration date. Where each date enters the analysis differs between the two designs and is specified below.

Death was ascertained using two data sources. First, deaths were identified within TBweb based on treatment outcomes recorded. Second, deaths were identified through probabilistic linkage with SIM, as described in Appendix §2. When date-of-death discrepancies between sources were identified, records were verified for individual identity and administrative consistency, and the earliest recorded date of death was used. Outcomes were classified as tuberculosis-attributable deaths and non-tuberculosis deaths. Tuberculosis-attributable deaths were defined using ICD-10 codes for underlying causes from SIM, supplemented by TBweb case-closure classifications. SIM records an underlying cause, defined as the condition that initiated the sequence of events leading to death, together with the contributing conditions entered on the remaining certificate lines. Deaths were counted as tuberculosis-attributable only where tuberculosis was the underlying cause; we repeated the cause attribution counting tuberculosis recorded on any line as a secondary analysis (Appendix §3.4). Non-tuberculosis deaths were defined as all other causes.

Covariates for all models were age group, sex, race, education, HIV status, diabetes, alcohol use, drug use, incarceration, homelessness, hospitalization at diagnosis, clinical tuberculosis classification, DOT, and municipality of treatment classified according to the Brazilian Institute of Geography and Statistics (IBGE) rural–urban typology (15), custody settings being folded into the urban reference level because that category is collinear with incarceration. All were defined at the time of treatment initiation, as TBweb records these characteristics at notification and does not reliably update them during treatment. Hospitalization at diagnosis is not routine in Brazil: the main indications are clinical severity, management of comorbidity including advanced HIV disease, and social circumstances in which outpatient treatment is not feasible. DOT is recommended for everyone treated for tuberculosis in Brazil but is not always achievable, for example where an individual lives far from the treating facility, where work schedules conflict with daily observation, or where housing is unstable.

## Statistical Analysis

### Multiple imputation

Missing data were handled using multiple imputation by chained equations with random-forest predictors, assuming the data were missing at random (16). The imputation model included all baseline covariates, the exposure, event indicators, and follow-up times, along with three predictors of missingness not in the adjustment set: calendar year of treatment start, recorded mental health condition, and setting of diagnosis. Missingness was related to recorded characteristics, most strongly to calendar year of treatment start and, for education, to social circumstances. The imputation model therefore conditioned on these along with the complete outcome and follow-up time (Appendix §4.1). Imputation was applied to covariates only. We generated five imputed datasets. Multivariable analyses were fitted separately in each imputed dataset and pooled using Rubin’s rules. Complete-case analyses were conducted as sensitivity analyses (Appendix §4.2).

### Mortality within the LTFU cohort

Among individuals who experienced LTFU, we estimated cumulative mortality using Kaplan–Meier analysis, with time measured from the declaration date, stratified by baseline characteristics and by the month of therapy at which disengagement occurred.

### Causal analyses: target trial emulation and rolling landmark

The primary analysis emulated a target trial from the start of treatment (17, 18), contrasting two strategies: disengagement from treatment at any point during the first six months, and remaining engaged throughout that window. Because an individual’s future disengagement cannot be known at treatment start, each individual was cloned into both strategies. Each clone was censored at the point at which the observed history diverged from its assigned strategy, and the resulting informative censoring was corrected with inverse-probability-of-censoring weights (19) estimated from the covariates listed above. An individual who was alive and still engaged within the window was compatible with both strategies, so deaths occurring at that time point contributed to both arms; surviving to the point of disengagement was not a condition of entering the analysis. Standardized cumulative mortality risk was computed under each strategy and contrasted at 60 months as a risk difference and a risk ratio. Follow-up was not extended beyond 60 months because the proportion of the cohort with sufficient follow-up declines steeply thereafter.

The timing of disengagement was addressed with a separate rolling landmark design (20). One landmark trial was opened for each day on which any individual was declared LTFU: individuals declared on that day formed the exposed arm, individuals still in care on that day formed the comparator, and follow-up for both arms began on that day. The 30-day grace period required by the LTFU definition therefore applies symmetrically, since both arms must be alive and event-free at the landmark. Trials were stacked and fitted with a single Cox model adjusted for the covariates listed above, with standard errors clustered on individual because individuals contribute to more than one trial, and absolute risks were obtained by g-formula standardization (21) to the disengaging population. Because the proportional-hazards assumption was rejected over a five-year horizon, the exposure was given a separate coefficient in each of five disjoint follow-up periods and the cumulative hazard assembled piecewise, so that the standardized risks do not assume a constant hazard ratio.

The two designs use different time origins (treatment start for the target trial emulation, the individual’s own declaration date for the landmark) and their magnitudes are therefore not comparable. The landmark exposed arm is smaller than the cohort total lost to follow-up because exposure must be indexed on a day of disengagement within the treatment course (Appendix §1). Both were repeated within subgroups defined by age group, sex, HIV status, homelessness, drug resistance and calendar period of treatment initiation. The two designs, and the time origin each uses, are shown in Figure 1. Both designs identify a causal effect only under assumptions the data cannot verify: that the arms are exchangeable conditional on the measured baseline covariates, so that no unmeasured determinant of both disengagement and death remains; that censoring at divergence from an assigned strategy is uninformative given those covariates and the weight model; and that both strategies were possible for every individual at the time origin. Unmeasured severity is the assumption most likely to fail, and Appendix §6 quantifies how strong such a confounder would need to be to explain the estimate away.

**Figure 1.**
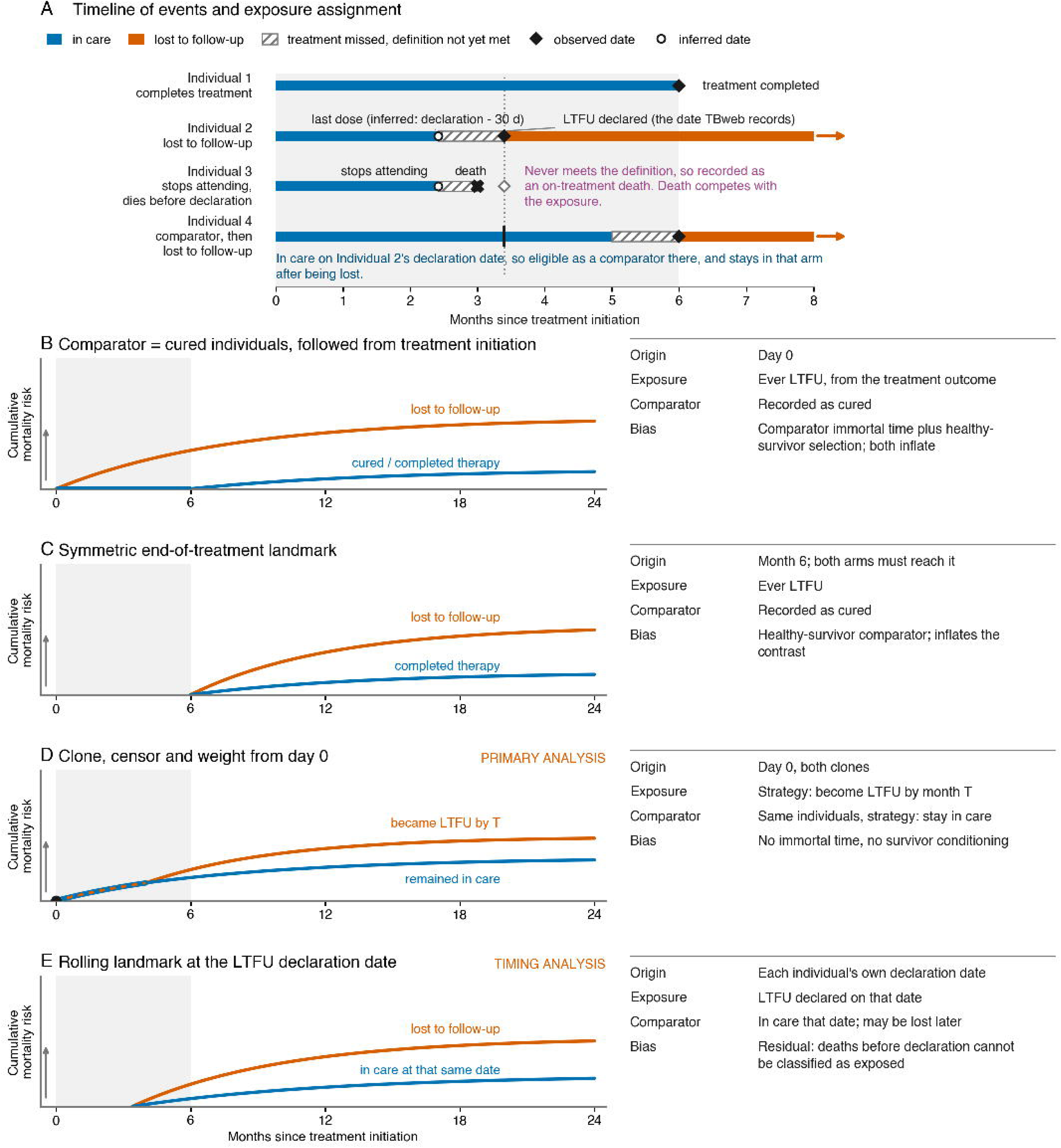
Time origins and comparators for estimating the effect of loss to follow-up on mortality. (A) Timeline of events and exposure assignment for four illustrative individuals: one who completes treatment; one lost to follow-up, showing the inferred date of last dose and the declaration date 30 days later; one who stops attending but dies before the definition is met, and is therefore recorded as an on-treatment death; and one who is in care on another individual’s declaration date, is eligible as a comparator there, and remains in the comparator arm after later being lost. (B–E) Four designs, each with its time origin, exposure definition, comparator and resulting bias. (B) Comparator of individuals recorded as cured, followed from treatment initiation, and (C) the same comparator under an end-of-treatment landmark. (D) Clone-censor-weight from day 0, the primary analysis, in which deaths occurring before the strategies diverge are counted in both arms. (E) Rolling landmark at each individual’s own declaration date. Curves are schematic and show the direction of each bias rather than fitted estimates.

### Cause-specific analyses

To assess whether the association between LTFU and mortality is consistent with the effects of treatment interruption or reflects residual confounding, we repeated the rolling landmark cohort analysis using cause-specific mortality outcomes, evaluating deaths due to tuberculosis and other causes. We utilized this design as the question concerns deaths after LTFU, whereas the target trial emulation shares deaths occurring before the two strategies diverge and dilutes the cause-specific contrast. Cause-specific Cox models were fitted by censoring deaths from the alternate cause at the time of occurrence. Cumulative incidence functions were then constructed (Aalen–Johansen) so that the tuberculosis and non-tuberculosis components sum to the all-cause risk difference. Deaths whose cause could not be classified were included in the non-tuberculosis class.

## Sensitivity analyses

We conducted sensitivity analyses to assess the robustness of the primary findings. First, cause-specific analyses were restricted to deaths with an ICD-10 classification from SIM only (Appendix §3.2), and were repeated counting tuberculosis recorded on any line of the death certificate rather than only as the underlying cause (Appendix §3.4). Second, analyses were stratified by calendar period (Appendix §5). Third, we repeated the primary rolling landmark cohort analysis in the complete-case set, restricted to individuals with complete data for all covariates (Appendix §4.3). Fourth, we quantified sensitivity to unmeasured confounding by computing an E-value for the primary risk ratio, which gives the minimum strength of association an unmeasured confounder would need with both disengagement and mortality to explain the estimate away (Appendix §6). In addition, we conducted descriptive supplementary analyses of temporal trends in LTFU incidence and post-LTFU outcomes across the study period. Finally, to evaluate how much of the variation in published estimates reflects study design rather than the underlying association, we applied the design used by most prior studies to this cohort, with follow-up from treatment initiation and a comparator of individuals recorded as cured, holding the covariates and horizon fixed. Because prior studies report relative measures, that comparison is made on the hazard-ratio scale.

### Software and reproducibility

Cohort construction and data management were performed in Python 3.13, and statistical analyses were conducted in R 4.5. Code and a reproducible analysis pipeline are available on https://github.com/evelepka/ltfu-outcomes-causal-inference.

### Ethics

Written permission for secondary use of surveillance data was obtained from the São Paulo State Health Department, and the study was approved by the Research Ethics Committee of the Instituto de Infectologia Emílio Ribas, São Paulo, Brazil.

## Results

### Study population

A total of 235,629 tuberculosis notifications were recorded in TBweb between 2013 and 2023, representing 219,282 individuals with a first tuberculosis episode. Of these, 6,552 were excluded due to a change of diagnosis, 12,430 due to missing or incomplete outcome information, and 1,801 due to transfer during treatment to another state or country. Further exclusions included individuals aged <15 years (n = 6,238), cases with information inconsistencies (n = 111), and deaths recorded on or before treatment initiation (n = 2,063).

Episodes with recorded outcome dates outside the study period (2013–2023) were also excluded (n = 17,624), and 21 individuals whose tuberculosis was identified only post-mortem were excluded as never at risk for loss to follow-up. Finally, 1,394 cases without a recorded treatment start date (789 LTFU, 605 continued-in-care) were excluded from the primary analysis to ensure consistent time-on-treatment alignment. The final primary cohort comprised 171,048 individuals. Of these, 20,830 (12.2%) experienced LTFU during their first episode. Among individuals who were not LTFU, 135,699 (90.3%) were programmatically classified as cured, 12,336 (8.2%) as deaths (5,520 tuberculosis-related and 6,816 non-tuberculosis-related), 1,044 (0.7%) as treatment failure, and 1,139 (0.8%) as other outcomes, including regimen changes due to toxicity.

Compared with individuals who continued in care, those who were LTFU were younger (median age 33 years [IQR 25–42] vs. 37 [IQR 27–51]), more frequently male (78.7% vs. 70.4%), and reported higher prevalence of homelessness (10.6% vs. 2.3%), alcohol use (29.9% vs. 16.2%), drug use (35.6% vs. 13.9%), and HIV (11.4% vs. 7.6%). They were also more often hospitalized at diagnosis (32.9% vs. 23.6%) and less likely to receive directly observed therapy (65.4% vs. 74.4%). (Table 1)

**Table 1.** Baseline characteristics of the study cohort, by loss-to-follow-up (LTFU) status (N = 171,048).

| **Characteristic** | **Continued in care**<br>**(N = 150,218)** | **LTFU**<br>**(N = 20,830)** |
| --- | --- | --- |
| *Sociodemographic* |  |  |
| Age, years, median (IQR) | 37 (27–51) | 33 (25–42) |
| Age group |  |  |
| 15–24 | 29,110 (19.4%) | 4,805 (23.1%) |
| 25–44 | 67,204 (44.7%) | 11,728 (56.3%) |
| 45–64 | 40,805 (27.2%) | 3,735 (17.9%) |
| ≥65 years | 13,099 (8.7%) | 562 (2.7%) |
| Male sex | 105,806 (70.4%) | 16,398 (78.7%) |
| Race/ethnicity |  |  |
| Black or Mixed | 68,084 (45.3%) | 10,246 (49.2%) |
| White | 61,908 (41.2%) | 6,356 (30.5%) |
| Indigenous and Asian | 1,637 (1.1%) | 146 (0.7%) |
| Educational attainment |  |  |
| ≤7 years | 48,026 (32.0%) | 7,107 (34.1%) |
| 8–11 years | 45,736 (30.4%) | 6,396 (30.7%) |
| ≥12 years | 16,168 (10.8%) | 1,327 (6.4%) |
| *Clinical comorbidities* |  |  |
| HIV/AIDS | 11,370 (7.6%) | 2,383 (11.4%) |
| Diabetes | 11,490 (7.6%) | 803 (3.9%) |
| Other immunosuppression | 2,038 (1.4%) | 112 (0.5%) |
| Mental health diagnosis | 2,326 (1.5%) | 296 (1.4%) |
| *Behavioral factors* |  |  |
| Alcohol use | 24,278 (16.2%) | 6,232 (29.9%) |
| Drug use | 20,874 (13.9%) | 7,421 (35.6%) |
| Tobacco use | 32,778 (21.8%) | 6,618 (31.8%) |
| *Social vulnerabilities* |  |  |
| History of incarceration | 19,282 (12.8%) | 1,394 (6.7%) |
| Homelessness | 3,440 (2.3%) | 2,206 (10.6%) |
| *Disease and treatment characteristics* |  |  |
| **Diagnosis setting** |  |  |
| Outpatient | 75,355 (50.2%) | 9,636 (46.3%) |
| Emergency department or inpatient | 56,237 (37.4%) | 8,768 (42.1%) |
| Institution-based active case-finding | 8,064 (5.4%) | 883 (4.2%) |
| Contact investigation | 3,644 (2.4%) | 412 (2.0%) |
| Community-based active case-finding | 3,321 (2.2%) | 550 (2.6%) |
| Inpatient admission at diagnosis | 35,413 (23.6%) | 6,855 (32.9%) |
| Laboratory-confirmed TB | 117,148 (78.0%) | 17,210 (82.6%) |
| Anatomical site of disease |  |  |
| Pulmonary | 121,957 (81.2%) | 18,099 (86.9%) |
| Extrapulmonary | 23,127 (15.4%) | 2,108 (10.1%) |
| Pulmonary plus<br>extrapulmonary/disseminated | 5,133 (3.4%) | 623 (3.0%) |
| Missing | 1 (0.0%) | 0 (0.0%) |
| Directly observed therapy (DOT) | 111,697 (74.4%) | 13,632 (65.4%) |

Percentages are column-wise within each group. Continuous age summarized as median (interquartile range).

Annual LTFU risk rose from approximately 10% in 2013–2018 to approximately 15% in 2020–2022, coinciding with the COVID-19 pandemic. Drug-susceptibility testing yielded a conclusive result in 21.7% of the cohort (n = 37,190/171,048). Among those tested, 1.51% (n = 562) had rifampin-resistant or multidrug-resistant tuberculosis and 2.25% (n = 836) had isoniazid mono-resistant disease. LTFU rates were broadly similar across drug-resistance categories: 11.7% (95% CI 9.3–14.7) among individuals with rifampin- or multidrug-resistant TB, 13.5% (13.1–13.9) in the drug-sensitive group, and 11.8% (11.7–12.0) among those without a recorded DST result. LTFU occurred at a median of 3.88 months (IQR 2.10–5.42) from treatment start (Figure 2A).

**Figure 2.**
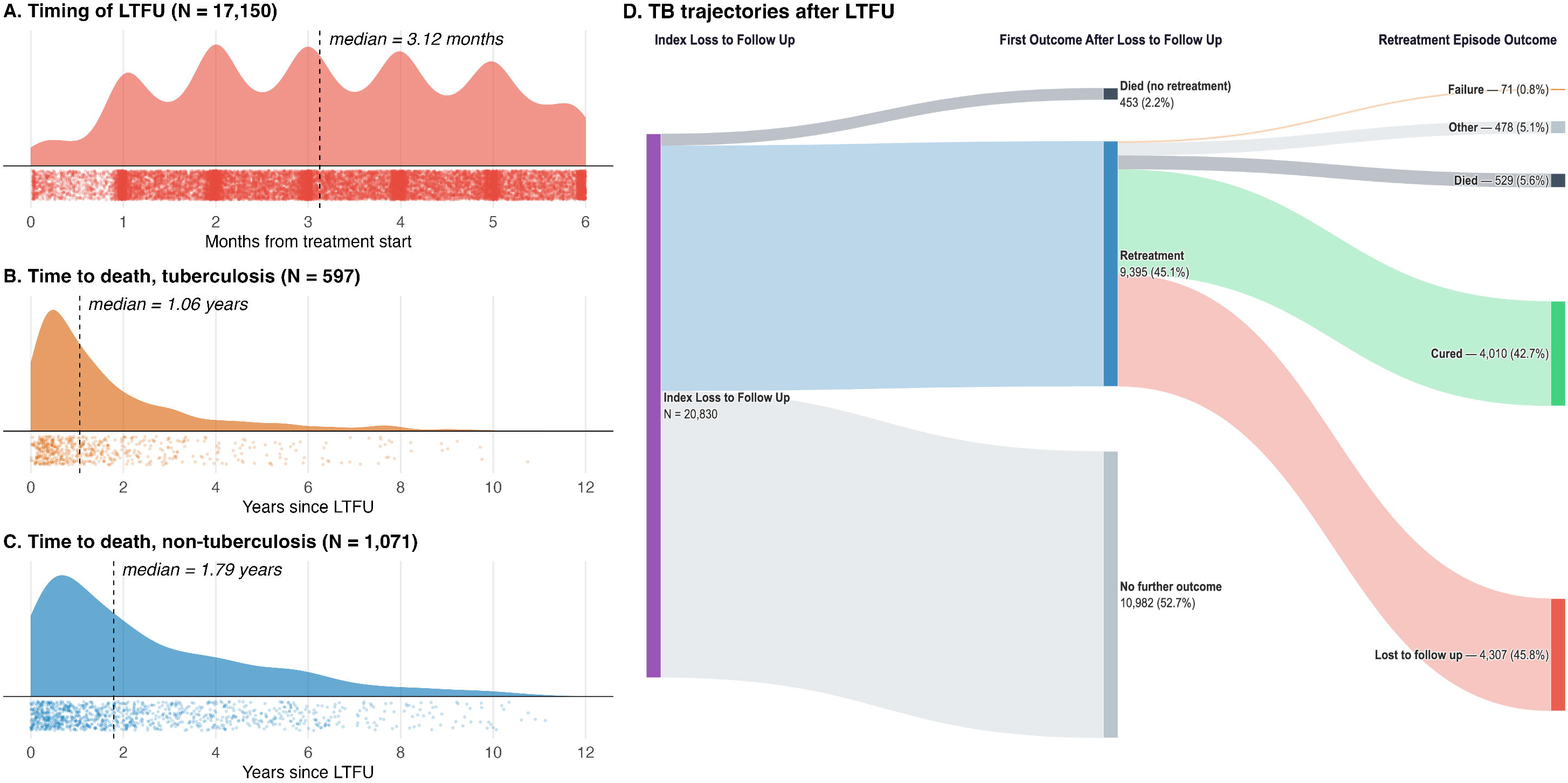
Timing and outcomes following loss to follow-up (LTFU). (A) Timing of LTFU from treatment initiation. (B) Time from LTFU to tuberculosis-attributed death (N = 597). (C) Time from LTFU to death not attributed to tuberculosis (N = 1,071). (D) Alluvial diagram showing post-LTFU trajectories (N = 20,830), from the first recorded outcome after LTFU through the outcome of a subsequent retreatment episode, when applicable. The diagram displays episode-level outcomes recorded in TBweb, including 453 deaths occurring before retreatment and 529 recorded as retreatment outcomes. These counts differ from the 1,668 deaths identified through linkage with the mortality registry, because the latter also capture deaths occurring after a retreatment episode had ended. All 1,668 were included in the analyses.

### Mortality after loss to follow-up

Among the 20,830 individuals who experienced LTFU, 9,395 (45.1%) subsequently began a retreatment episode, 453 (2.2%) died without returning to treatment, and 10,982 (52.7%) had no further recorded episode (Figure 2D). Among those who returned, 4,010 (42.7%) were cured, and 4,307 (45.8%) were again LTFU. Post-LTFU observation totaled 111,703 person-years, with a median of 5.0 years (IQR 2.5–8.0) per individual; 1,668 (8.0%) died, at a median of 17.5 months (IQR 6.7–38.9) after LTFU, a crude mortality rate of 14.9 deaths per 1,000 person-years (95% CI 14.2–15.7).

Crude five-year mortality after LTFU varied more than 5-fold across baseline characteristics (Figure 3): 20.0% among people living with HIV (PLHIV) compared with 6.3% among HIV-negative individuals; 17.5% among individuals hospitalized at diagnosis versus 3.0% among those who were not; 14.5% among individuals experiencing homelessness compared with 7.1% among housed individuals; 11.0% among those reporting alcohol use versus 6.6% among those not; and rising with age from 4.4% at 15–24 years to 16.4% at 65 years or above. Mortality was highest among individuals disengaging earliest, falling from 12.3% among those who disengaged in the first month of therapy to 4.1% in the sixth.

**Figure 3.**
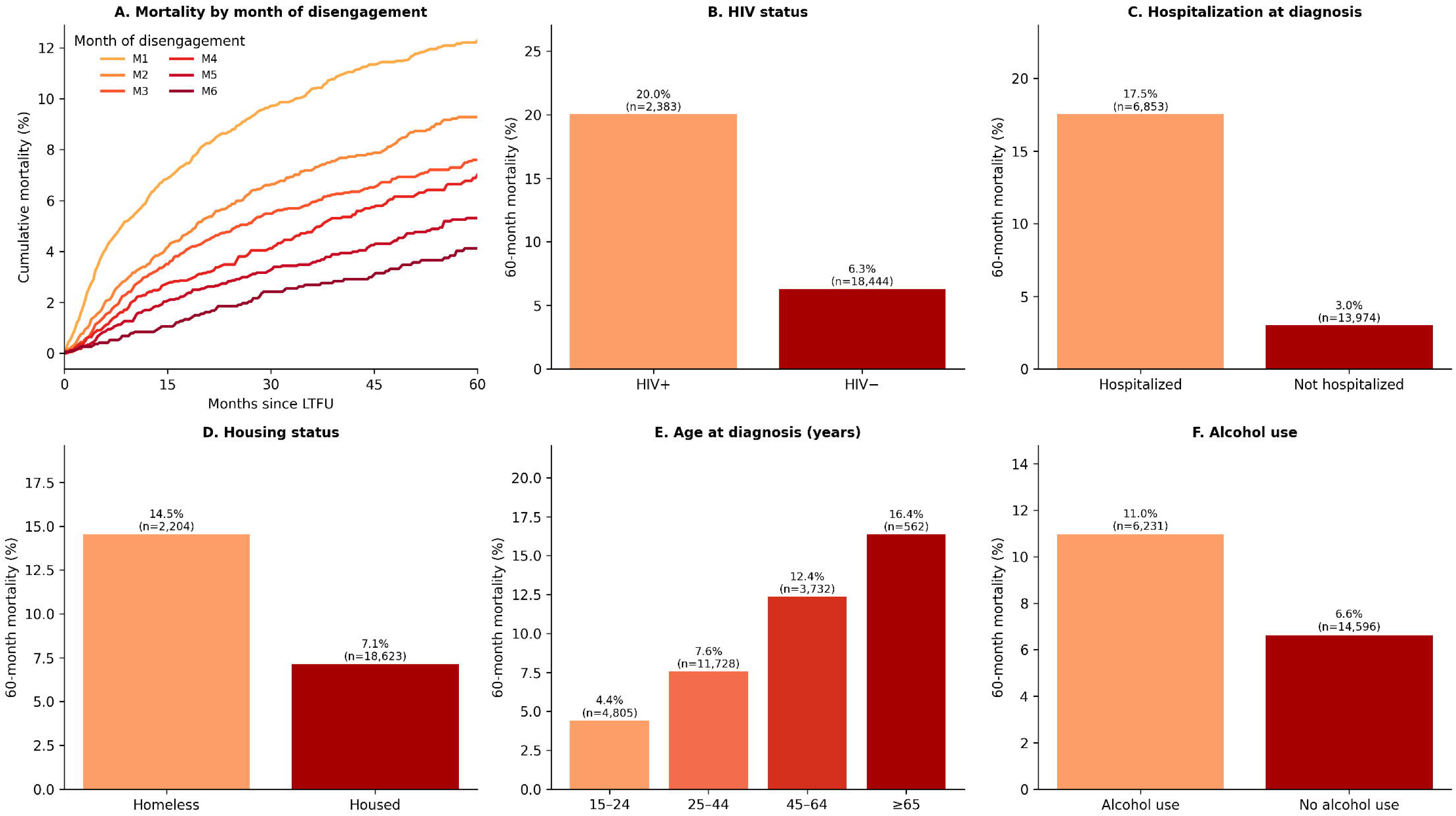
Cumulative mortality among individuals lost to follow-up. (A) Cumulative mortality by month of disengagement, within the LTFU cohort. (B–F) Five-year cumulative mortality stratified by HIV status (B), hospitalization at diagnosis (C), housing status (D), age group (E), and alcohol use (F). Crude Kaplan–Meier estimates with time measured from LTFU. Month of disengagement m refers to LTFU occurring anytime within that month (e.g. LTFU in month 6 refers to disengagement between months 5 and 6).

### Causal effect of LTFU on mortality

The primary analysis emulated a target trial from the start of treatment, contrasting disengagement at any point during the first six months with continued engagement. At 60 months, standardized mortality risk was 11.62% under the disengagement strategy against 9.40% under continued engagement, an absolute risk difference of 2.23 percentage points (95% CI 1.88–2.51) and a risk ratio of 1.24 (95% CI 1.20–1.27) (Figure 4).

**Figure 4.**
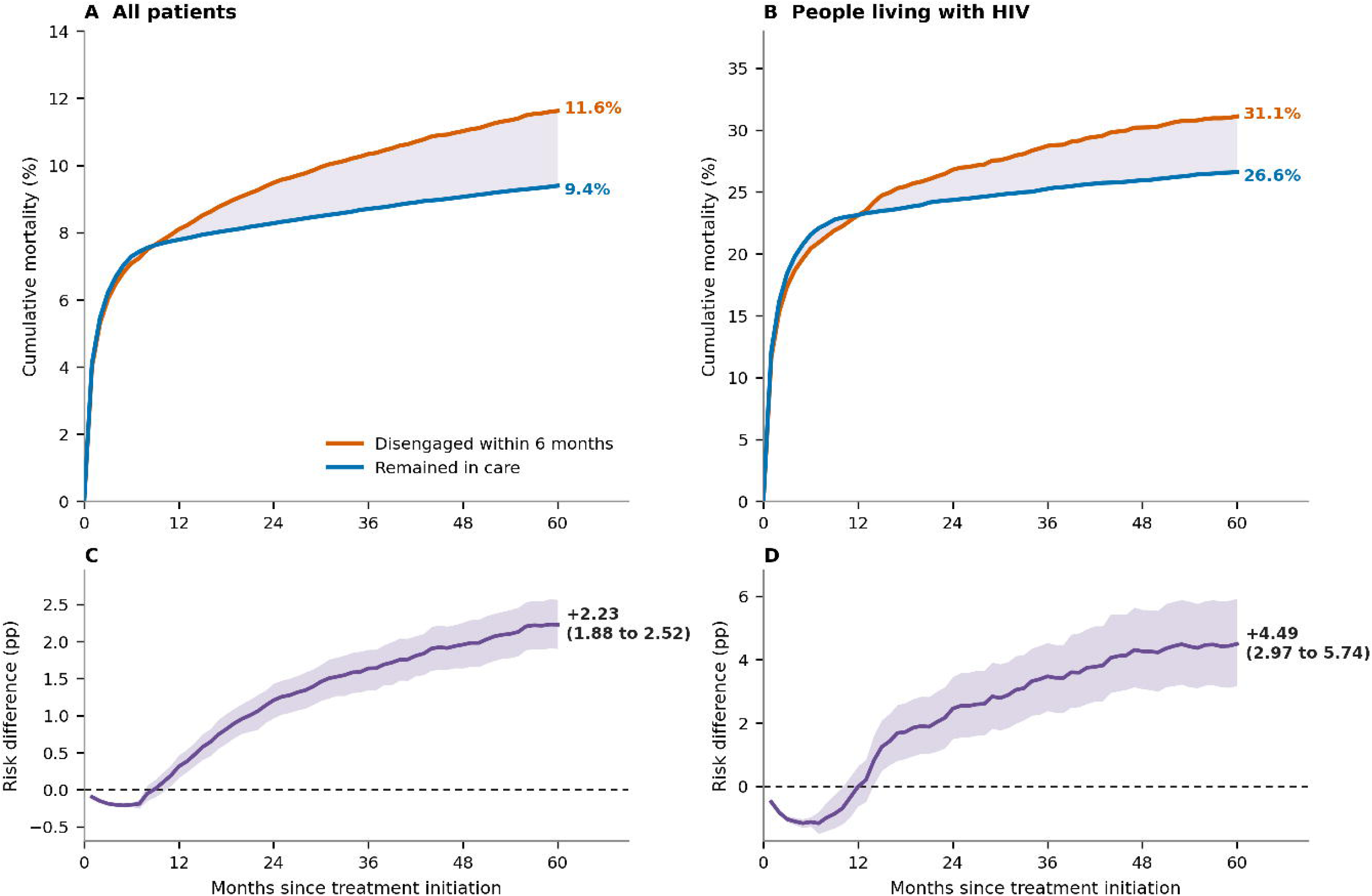
Standardized cumulative mortality under each assigned strategy. (A) All individuals and (B) people living with HIV: weighted, model-standardized cumulative mortality under disengagement from treatment within the first six months (orange) and under continued engagement (blue), from the clone-censor-weight target trial emulation with time measured from treatment start. (C, D) The corresponding risk difference with a 95% patient-level bootstrap band (B = 300). Risk differences below zero in the early months reflect the exposure definition rather than benefit, since an individual who dies early cannot be recorded as lost to follow-up and those deaths can only be classified as occurring in care.

In the rolling landmark analysis, the five-year standardized risk difference was similar for disengagement at any point in the first four months, from 3.10 percentage points (95% CI 1.83–4.35) in month 1 to 2.95 (2.23–3.99) in month 4, then fell to 2.21 (1.54–3.16) in month 5 and 1.04 (−0.01 to 2.16) in month 6 (Figure 5C).

**Figure 5.**
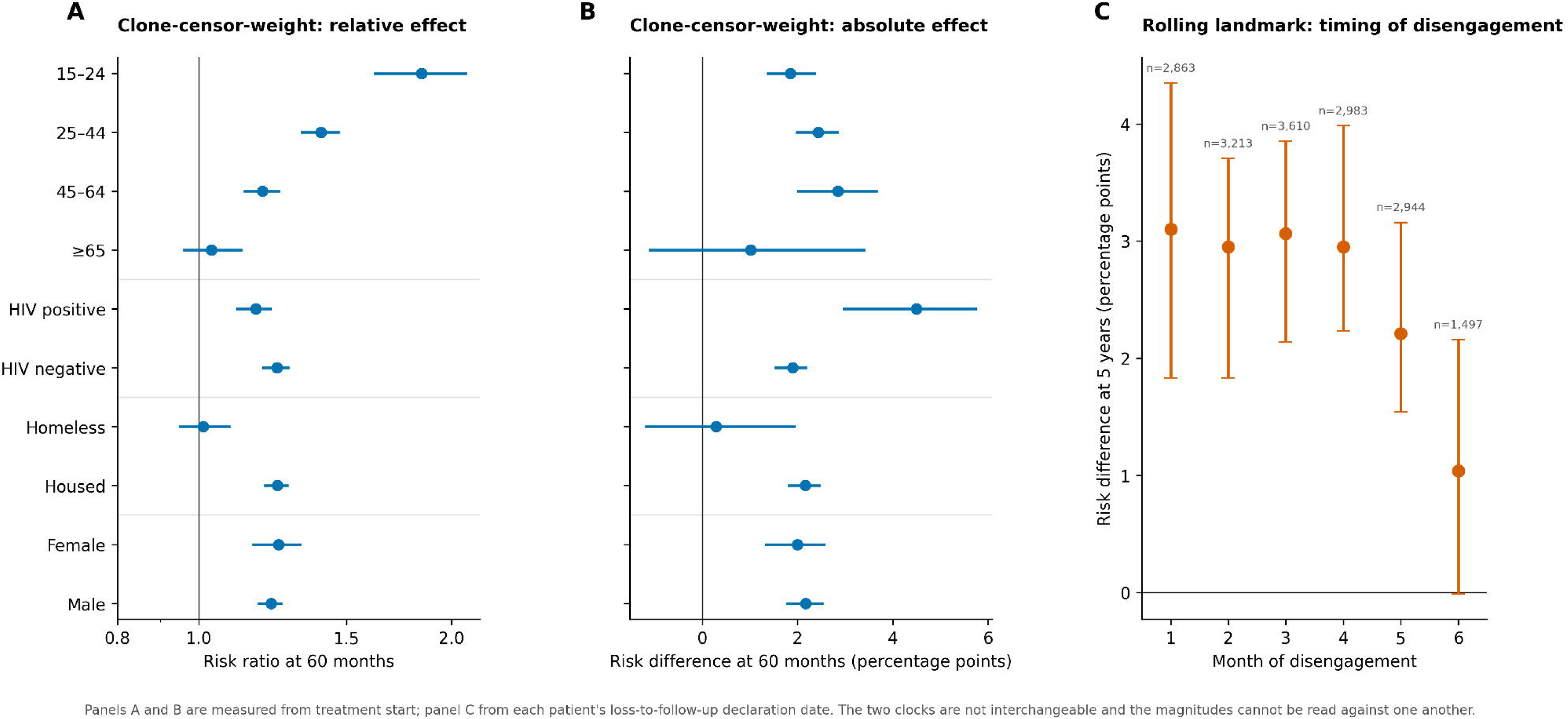
Causal effect of LTFU on mortality. (A) Risk ratio at 60 months by subgroup and (B) standardized absolute risk difference at 60 months by subgroup, both from the clone-censor-weight target trial emulation with time measured from treatment start. (C) Standardized absolute mortality risk difference at five years by month of disengagement, from the rolling landmark analysis. LTFU in month m refers to disengagement occurring anytime within that month (e.g. LTFU in month 6 refers to disengagement between months 5 and 6).

### Cause-specific evidence: TB-attributable vs. non-TB mortality

A cause of death was available from either source for 85.9% of deaths in the exposed arm and 88.3% in the comparator arm. A cause coded in the national mortality registry was available for 39.8% and 44.5%, respectively; the remainder was taken from the programmatic outcome recorded in TBweb. When both sources cited the same cause of death (n = 2,618), they agreed on tuberculosis versus non-tuberculosis attribution in 76.2% of cases (Cohen’s κ = 0.52).

The excess was concentrated in deaths attributed to tuberculosis. On the absolute scale, the five-year excess of 2.68 percentage points (95% CI 2.33 to 3.07) partitioned into 1.64 percentage points attributed to tuberculosis (1.37 to 1.89) and 1.04 percentage points not attributed to tuberculosis (0.73 to 1.39). Tuberculosis therefore accounted for 61% of the total excess. Restricting cause of death to ICD-10 codes recorded in the mortality registry gave the same ordering (Appendix §3.2). Counting tuberculosis recorded on any line of the death certificate rather than only as the underlying cause classified 26% more deaths as tuberculosis-related and raised the tuberculosis share of the excess from 61% to 63% (1.67 versus 1.64 percentage points), with the reclassified deaths falling in both arms (Appendix §3.4).

### Effect modification of LTFU

Effects were compared across subgroups within the same target trial emulation, at 60 months (Figure 5A and 5B). On the relative scale, the risk ratio was highest in the youngest individuals (15–24 years, 1.84, 95% CI 1.62–2.08) and declined with age, to 1.40 (1.33–1.47) at 25–44 years, 1.19 (1.14–1.25) at 45–64 years, and 1.04 (0.96–1.12) at 65 years or above. PLHIV had a lower risk ratio (1.17, 95% CI 1.11–1.22) than HIV-negative individuals (1.24, 1.19–1.28), and individuals experiencing homelessness a lower ratio (1.01, 0.95–1.09) than housed persons (1.24, 1.20–1.27). Estimates were similar by sex (female 1.25, 1.16–1.32; male 1.22, 1.18–1.25) and across calendar periods (2013–2019: 1.25, 1.20–1.30; 2020–2023: 1.21, 1.15–1.28).

On the absolute scale the ordering reverses. The 60-month risk difference was largest among PLHIV (4.49 percentage points, 95% CI 2.97–5.74), more than twice the figure in HIV-negative persons (1.90, 1.53–2.18), and rose with age from 1.85 (1.38–2.36) at 15–24 years to 2.85 (2.02–3.66) at 45–64 years. Among the small group with drug-resistant tuberculosis, it was 8.35 percentage points (1.82–15.25).

## Discussion

Loss to follow-up during tuberculosis treatment raised standardized five-year mortality by 2.23 percentage points, and the excess was concentrated in tuberculosis-attributable deaths. Five-year risk differences were similar for disengagement at any point during the first four months of treatment, attenuating thereafter. The relative effects of LTFU were largest among younger, stably housed individuals, in whom the competing-mortality baseline is low, whereas on the absolute scale the greatest LTFU-attributable mortality burden fell on PLHIV. These findings, in a setting with high DOT coverage, point to the need for more effective models of care engagement throughout the course of treatment, particularly among PLHIV.

Historically, LTFU has been considered a negative programmatic indicator for tuberculosis treatment programs, presumed to lead to negative health outcomes; however, its effects on mortality have not been well characterized. As LTFU has been increasing in Brazil in recent years, alongside tuberculosis mortality, it is critical to understand and quantify the relationship between this process indicator and health outcomes. Quantifying the impacts of LTFU on mortality, including its effects by timing of disengagement and risk group, will be important towards developing interventions that can be tested to address this. Treatment observation has been the most widely implemented strategy for tuberculosis treatment adherence (22); however, given that more than 65% of individuals who were LTFU in this study were receiving DOT, it is clear that additional approaches are needed.

Prior studies have found that LTFU during tuberculosis treatment increases mortality (6–11), but their estimates may be considerably inflated due to the choice of the comparator. Adjusted hazard ratios from the studies that compared individuals lost to follow-up with other patients under treatment range from 3.65 to 8.9 (8–10), and the studies that report standardized mortality ratios by treatment outcome find them two to seven times higher after loss to follow-up than after cure or treatment success (6, 11). Comparing individuals lost to follow-up with those who completed treatment contrasts them with a group defined by a favorable future outcome, since completion requires survival, continued care, and response to therapy (6, 7, 9, 11). In our own data, holding the cohort, covariates and horizon fixed and changing only the design, a comparator of individuals recorded as cured gave a five-year adjusted hazard ratio of 3.20 (95% CI 2.97–3.45), compared with 1.58 (1.48–1.70) for the design reported here. The choice of the time origin also affects estimates. When follow-up is counted from treatment initiation, LTFU appears spuriously protective in early follow-up, because classification requires survival for at least 30 days post-disengagement (e.g. an individual who dies within 30 days of disengagement is recorded as an on-treatment death) (5). On the other hand, when follow-up is counted from the time when treatment should have been completed (e.g. 6 months), a healthy survivor bias is introduced in both arms.

The designs used here address these issues: the target trial emulation assigns strategies at treatment start, before disengagement can be known; the rolling landmark analysis begins follow-up at each individual’s own declaration date, so no time at risk precedes the exposure; and in both designs, the comparator is individuals still in treatment at the time origin rather than individuals who completed therapy. However, residual survival and selection biases remain.

These biases are evident in the risk difference below zero in early follow-up (Figure 4), which does not reflect a protective effect of disengagement. Becoming LTFU requires surviving long enough to be declared lost to follow-up, 30 days beyond the last recorded contact.

We found a modest concentration of excess mortality in tuberculosis-attributable rather than non-tuberculosis deaths. However, distinguishing the two can be challenging. People who complete tuberculosis treatment continue to die at several times the rate of the general population: a pooled standardized mortality ratio of 2.91 (95% CI 2.21–3.84), rising to 3.76 (3.04–4.66) when restricted to those with confirmed cure or completion, with cardiovascular disease the single largest cause of post-treatment death (20%, 15–26) rather than tuberculosis itself (23). In Brazil, the excess in deaths from natural causes other than tuberculosis persists 14 years after treatment completion (24), and in São Paulo State 20% of deaths within five years of diagnosis had tuberculosis as the underlying cause, with a further 14% recording it elsewhere on the certificate (6). Both compare people with tuberculosis against general or unexposed populations, so they establish that mortality following tuberculosis is broad and prolonged without quantifying how much of it disengagement causes. A broader rule does not resolve this: counting tuberculosis wherever it appears on the certificate raised the tuberculosis share of the excess only from 61% to 63%.

Social and structural disadvantage lies upstream of both tuberculosis disease and LTFU (25, 26), and tuberculosis itself lies on the pathway from disadvantage to death. The disproportionate mortality burden among people experiencing homelessness (27) and PLHIV reflects this. Our analyses target the effect of LTFU given the social and clinical characteristics present at treatment initiation, so LTFU is best understood as one modifiable step along that pathway rather than a cause separable from its social determinants. Within that framing, disengagement carries a measurable mortality cost of its own. Addressing its structural drivers, including housing instability, substance use (28), mental-health needs (29) and HIV co-infection, is therefore central to reducing both the incidence of LTFU and the mortality burden it imposes.

This analysis has several limitations. Residual confounding remains possible because markers or determinants of disease severity, such as HIV viral load, CD4 count, and radiographic extent of disease, were unavailable in TBweb. We quantified the strength that an unmeasured confounder would need to explain away the primary estimate and found that a confounder of moderate strength could account for it. Because clinical characteristics are recorded at notification and not reliably updated, we could not adjust for changes arising after treatment start, so confounding by events during treatment, such as clinical deterioration, a new diagnosis, or a change in living circumstances, is not addressed. Mortality ascertainment relied in part on probabilistic linkage with SIM, which may have under-ascertained deaths among highly mobile populations. Cause of death could not be classified for 13.6% of deaths within five years among individuals lost to follow-up and 11.9% among those continuing in care, so the split between tuberculosis- and non-tuberculosis-attributable mortality remains subject to misclassification.

Cause was drawn from a death certificate for a similar share of tuberculosis deaths in each arm (34.4% and 34.3%), so differential attribution is unlikely to explain the tuberculosis-specific excess. The death-certificate only (SIM) sensitivity analysis guards against differential attribution, and tuberculosis continued to account for the majority of classifiable excess deaths (Appendix §3.2). We cannot fully exclude informative missingness within exposure- or return-status strata, although attribution availability did not differ materially across these strata. Finally, the date of disengagement is approximated from operational TBweb fields and could not be verified against direct adherence records.

LTFU during tuberculosis treatment raised standardized five-year mortality by 2.23 percentage points, one additional death for every 45 individuals who disengaged and one for every 22 among PLHIV. The excess was similar for disengagement at any point during the first four months of treatment, so there is no window in which interruption is safe. This occurred in a program with high DOT coverage, so sustaining engagement requires more than observing individual doses. Preventing LTFU by addressing the structural and clinical vulnerabilities that increase the risk of disengagement, including housing instability, substance use, and HIV co-infection, and provision of person-centric, integrated models of care, should be programmatic priorities.

## Data Availability

All data underlying the findings of this study are available in a public GitHub repository at https://github.com/evelepka/ltfu-minimal-dataset

## Acknowledgments

The authors received no specific funding for this work.

## Supporting information captions

**S1 Appendix.** Supplementary methods, tables, and figures, including cohort definition and exclusion-criterion sensitivity, record linkage between the surveillance and mortality registries, cause-of-death attribution sensitivity analyses, missing-data and complete-case analyses, calendar-period analyses, E-value sensitivity to unmeasured confounding, and a summary table of estimands, comparators and estimates.

**S1 Checklist.** STROBE statement checklist for cohort studies.

