## Appendix for "Causal effect of loss to follow-up on mortality in a population-based tuberculosis cohort in Brazil"

**Contents**

1. Cohort definition and exclusion-criterion sensitivity

2. Record linkage between TBweb and the mortality registry

3. Cause-of-death attribution sensitivity

4. Missing data and complete-case sensitivity

5. Calendar-period analyses

6. Sensitivity to unmeasured confounding (E-value)

7. Summary of estimands and estimates

### 1. Cohort definition and exclusion-criterion sensitivity

#### 1.1 Cohort attrition flow

The cohort was constructed from 235,629 TBweb notifications (219,282 individuals with a first tuberculosis episode). Exclusions: 6,552 change-of-diagnosis cases; 12,430 with missing or incomplete outcome; 1,801 inter-state transfers during treatment; 6,238 aged under 15 years; 111 data inconsistencies; 2,063 deaths on or before treatment initiation; 17,624 with treatment end dates outside the study period; 21 individuals identified only post-mortem (never at risk for LTFU); and 1,394 without recorded treatment start. Final cohort: 171,048 individuals (Figure S1) (20,830 LTFU, 12.2%; 150,218 continued in care). An unrecorded programmatic outcome could in principle mask undocumented disengagement, but of the 12,430 excluded on that ground, only 6 had both a valid in-window treatment-closure date and a recorded treatment start date, and only 2 met every remaining eligibility criterion. Re-classifying that group as lost to follow-up would add 2 individuals to the cohort and 2 to the exposed arm, so it would not alter any estimate.

The two designs use the same cohort of 171,048 individuals but different exposed sets. The target trial emulation assigns every individual to both strategies at treatment start, so all 20,830 who were lost to follow-up contribute. The rolling landmark indexes exposure on the day of disengagement, which requires a day that exists on the 1 to 180 axis of therapy received, so three groups cannot enter: 377 with a programmatic outcome of primary abandonment, who were notified but never established on treatment and are analysed separately; 1,169 whose case closure was recorded less than 31 days after treatment start, so that subtracting the 30-day declaration window leaves no positive day; and 2,172 who disengaged after day 180, beyond the intended course. The landmark exposed arm therefore comprises 17,112 individuals. The excluded groups remain in the comparator-eligible pool for as long as they are in care.

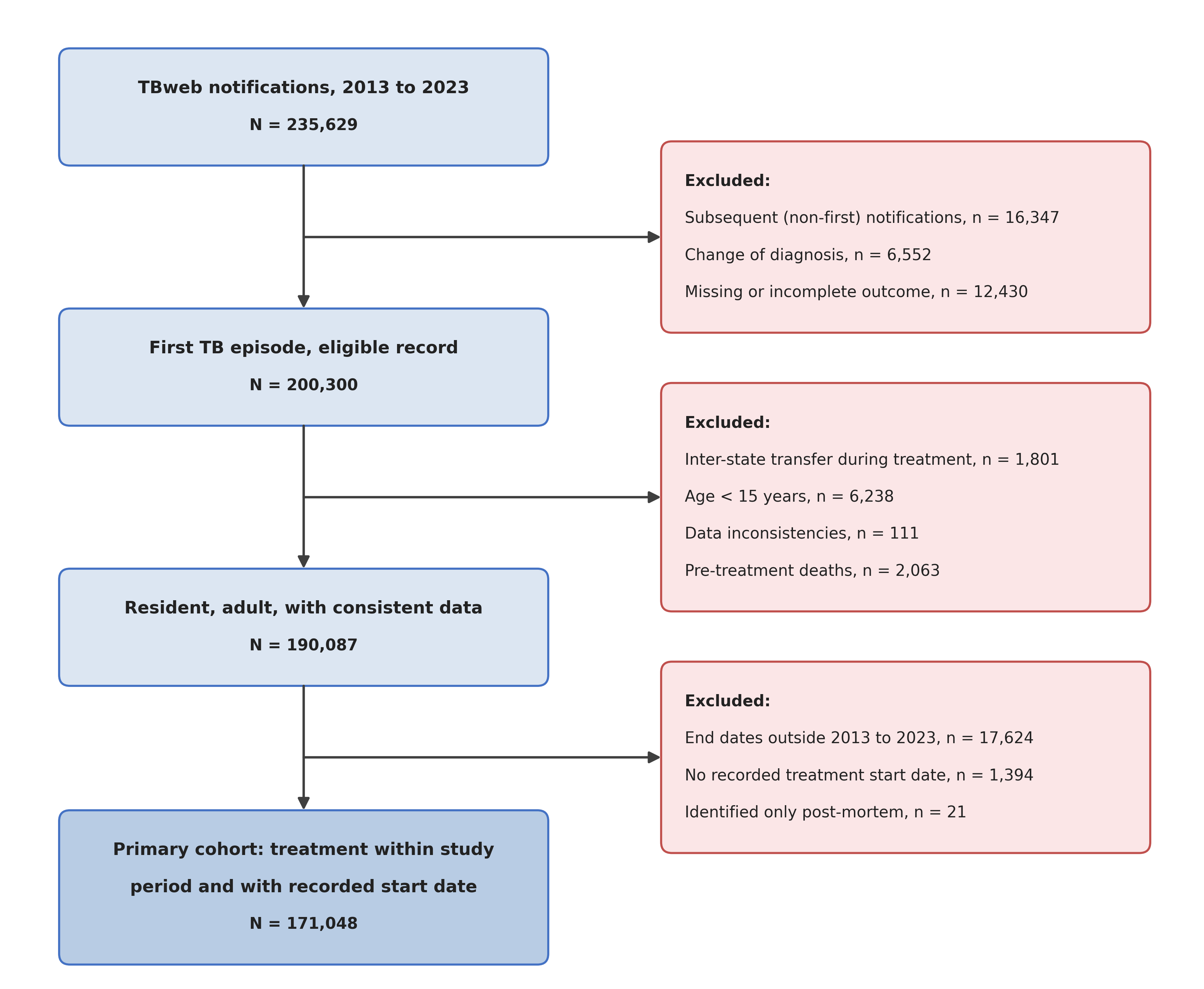

**Figure S1. CONSORT Diagram**.

### 2. Record linkage between TBweb and the mortality registry

Mortality was ascertained by probabilistic record linkage between tuberculosis notifications reported in São Paulo State between 2013 and 2024 and death records from the national Mortality Information System (SIM) for the same period. Records from TBweb (n = 249,208) and SIM were compared on the individual’s name, the mother’s name and the date of birth; similarity scores were computed for candidate pairs and blocking was applied to limit false positives. The automated step returned 12,813 candidate pairs. Each was reviewed manually, and pairs with discrepancies, incomplete fields, or non-informative entries such as “unknown” or “ignored” were rejected, leaving 10,056 high-confidence matches. All personal identifiers were removed after linkage.

A linkage that failed more often in one arm than the other would bias the cause-specific contrast, so we compared ascertainment by exposure status. That comparison is reported with the cause-of-death attribution below.

### 3. Cause-of-death attribution sensitivity

#### 3.1 Underlying-cause attribution

Each death was classified as TB-attributable if either (i) the linked death certificate from the national Mortality Information System (SIM) carried an ICD-10 underlying-cause code for tuberculosis (A15–A19), tuberculosis sequela (B90), or HIV disease with tuberculosis (B200); or (ii) no death-certificate code was available and the death was recorded as a TB death in the State TB notification registry (TBweb). Deaths were classified as non-TB when the death certificate carried a non-TB code or the registry recorded a non-TB death. Respiratory deaths (ICD-10 J-codes) and other HIV-related deaths (B20–B24 except B200) were treated as of unknown cause under this primary, “hybrid” attribution.

#### 3.2 Death-certificate–only sensitivity

To evaluate the influence of misclassification in the registry-recorded cause of death, we repeated the cause-specific analyses restricted to deaths carrying a death-certificate (SIM) ICD-10 code. Over five years the risk difference for deaths attributed to tuberculosis was 0.51 percentage points (95% CI 0.36 to 0.60), and for deaths attributed to other causes 0.36 (0.20 to 0.63). In this analysis tuberculosis continued to account for the majority of classifiable excess deaths, so the concentration in tuberculosis deaths does not depend on the programmatic outcome codes used to supplement registry causes.

#### 3.3 Attribution availability by exposure arm

A cause of death was available from either source for 85.9% of deaths in the LTFU arm and 88.3% in the comparator arm. A cause coded in the mortality registry was available for 39.8% and 44.5% respectively. Availability differs little between arms, and the difference that does exist acts against the concentration in tuberculosis rather than in favor of it, because deaths carrying no specific cause fall into the residual class. Tuberculosis deaths were ascertained mainly through TBweb in both arms (95% and 96%), whereas non-tuberculosis deaths were ascertained through TBweb and the registry in similar proportions (58% and 49% among individuals lost to follow-up; 56% and 53% among those continuing in care).

We additionally quantified concordance between the two cause-of-death sources directly. Among the 2,618 cohort deaths for which both a TBweb treatment-outcome death classification (tuberculosis-attributable death vs non-tuberculosis death) and a SIM ICD-10 underlying cause were available, the sources agreed on tuberculosis versus non- tuberculosis attribution in 76.2% of cases (Cohen’s κ = 0.52, moderate agreement); taking SIM as the reference, the TBweb classification had a sensitivity of 73.3% and a specificity of 78.9% for a tuberculosis -coded underlying cause. A SIM ICD-10 code was available for only 6,200 of the 12,336 cohort deaths carrying a TBweb death classification, which is why the primary (hybrid) attribution supplements SIM with TBweb case-closure codes. This differs from Ranzani et al. (*Lancet Infect Dis*, 2020), the São Paulo tuberculosis long-term mortality cohort, who relied on SIM ICD-10 coding with probabilistic linkage and manual record review and classified deaths through the WHO ICD-10 cause hierarchy. Our hybrid approach trades some uniformity of source for higher completeness of cause ascertainment in a registry-linked cohort in which roughly half of deaths lack an ICD-10 code. The moderate TBweb–SIM agreement and the incomplete ICD-10 coverage together motivate the death-certificate–only sensitivity analysis (§3.2).

#### 3.4 Tuberculosis recorded on any line of the death certificate

The attribution used in the main analysis is the underlying cause of death, the single condition the coder identifies as having initiated the sequence leading to death. Tuberculosis may instead be recorded as a contributing condition, in which case the underlying-cause rule does not count the death as tuberculosis-related. We therefore repeated the cause decomposition counting tuberculosis recorded anywhere on the certificate, which classified 26% more deaths as tuberculosis-related. At five years, the tuberculosis-attributable risk difference was 1.67 percentage points (95% CI 1.40 to 1.93) under this wider rule compared with 1.64 (1.37 to 1.89) under the underlying-cause rule, and the difference not attributed to tuberculosis was 1.00 compared with 1.04, so the tuberculosis share of the total excess rose from 61% to 63%. Almost all of the reclassified deaths came from the residual class whose cause could not be assigned to either category, whose contribution fell from 0.35 to 0.31 percentage points. They fell in both arms: the five-year risk of a tuberculosis-attributed death rose from 2.83% to 3.01% among individuals lost to follow-up and from 1.19% to 1.33% among those continuing in care, so the relative effect is slightly smaller under the wider rule (risk ratio 2.26 versus 2.37).

**Table S1. Characteristics at diagnosis of deaths among individuals lost to follow-up, by cause class.** Percentages are of each cause class, so each variable sums to 100 within a column. The not-tuberculosis class is the residual, all deaths minus those attributed to tuberculosis, and therefore includes deaths whose recorded cause was not specific enough to classify.

| **Characteristic** | **Level** | **Not tuberculosis, n (%)** | **Tuberculosis, n (%)** |
| --- | --- | --- | --- |
| Age group | 15-24 | 134 (12.5) | 82 (13.7) |
|  | 25-44 | 563 (52.6) | 333 (55.8) |
|  | 45-64 | 307 (28.7) | 161 (27.0) |
|  | 65+ | 67 (6.3) | 21 (3.5) |
| Sex | Female | 280 (26.1) | 112 (18.8) |
|  | Male | 791 (73.9) | 485 (81.2) |
| HIV status | Negative | 671 (62.7) | 514 (86.1) |
|  | Positive | 400 (37.3) | 83 (13.9) |
| Hospitalized at diagnosis | | 761 (71.1) | 480 (80.4) |
| Diabetes | | 63 (5.9) | 23 (3.9) |
| Alcohol use | | 406 (37.9) | 272 (45.6) |
| Drug use | | 405 (37.8) | 281 (47.1) |
| Tobacco use | | 288 (26.9) | 229 (38.4) |
| Mental health condition present | | 18 (1.7) | 12 (2.0) |
| Other immunosuppressive condition present | | 13 (1.2) | 2 (0.3) |
| Experiencing homelessness | | 195 (18.2) | 128 (21.4) |
| Incarcerated | | 22 (2.1) | 10 (1.7) |
| Clinical form | Extrapulmonary | 135 (12.6) | 33 (5.5) |
|  | Pulmonary | 861 (80.4) | 539 (90.3) |
|  | Pulmonary and Extrapulmonary or disseminated | 75 (7.0) | 25 (4.2) |
| Drug-resistance status | Not Evaluated | 796 (74.3) | 402 (67.3) |
|  | Resistant (Any) | 10 (0.9) | 10 (1.7) |
|  | Sensitive | 265 (24.7) | 185 (31.0) |

### 4. Missing data and complete-case sensitivity

#### 4.1 Missingness patterns

Four covariates were incomplete: education (27.1%), self-reported race (13.3%), DOT status (7.0%), and clinical form (one individual). Exposure, outcome, and follow-up time were complete. Missingness was more frequent in the LTFU arm for race (19.6% vs 12.4%), DOT status (10.0% vs 6.6%), and education (28.8% vs 26.8%). Race missingness increased from 10.6% of 2014 treatment starts to 31.1% in 2023. Education was unrecorded for 44.1% of people experiencing homelessness versus 26.5% housed, 40.9% with a recorded mental health condition, and 36.5% hospitalized at diagnosis.

Standardized over the 14 variables recorded for every individual, two-year mortality for unrecorded versus recorded values differed by −0.51 percentage points (95% CI −0.83 to −0.12) for race, 2.98 (2.69 to 3.28) for education, and 5.17 (4.59 to 5.73) for DOT status. The corresponding unstandardized differences were 0.01, 6.39, and 6.78 (Table S1). The contrast is taken at two years rather than five so that it can be estimated on the 161,922 individuals with a full horizon of potential follow-up.

Calendar year and mental health condition are among the strongest predictors of missingness, and both are included in the imputation model as auxiliary variables. Those without recorded DOT status had two-year mortality of 14.9%, closer to those not receiving DOT (13.9%) than to those receiving it (6.6%). As 78.8% of recorded values indicated DOT, imputation under missing at random may misclassify unrecorded status. DOT status is an adjustment covariate, so misclassification would leave that adjustment incomplete rather than over-corrected.

**Table S2. Per-variable baseline missingness in the primary cohort (N = 171,048).** The final column is the difference in two-year mortality between individuals whose value was not recorded and those whose value was, standardized over the 14 variables recorded for every individual, shown only where enough values are missing to estimate it.

| **Variable** | **N missing** | **% missing** | % missing, LTFU | % missing, in care | Standardized difference in 2-year mortality, unrecorded vs recorded (percentage points, 95% CI) |
| --- | --- | --- | --- | --- | --- |
| Age at TB diagnosis | 0 | 0.00% | — | — | — |
| Sex | 0 | 0.00% | — | — | — |
| Race | 22,671 | 13.25% | 19.60% | 12.37% | −0.51 (−0.83 to −0.12) |
| Education | 46,288 | 27.06% | 28.80% | 26.82% | 2.98 (2.69 to 3.28) |
| HIV/AIDS | 0 | 0.00% | — | — | — |
| Diabetes | 0 | 0.00% | — | — | — |
| Alcohol use | 0 | 0.00% | — | — | — |
| Drug use | 0 | 0.00% | — | — | — |
| Homelessness | 0 | 0.00% | — | — | — |
| Incarceration | 0 | 0.00% | — | — | — |
| Hospitalization at diagnosis | 0 | 0.00% | — | — | — |
| Clinical form | 1 | < 0.01% | — | — | — |
| DOT status | 11,914 | 6.97% | 9.99% | 6.55% | 5.17 (4.59 to 5.73) |
| Mental health | 0 | 0.00% | — | — | — |
| Other immunocompromising condition | 0 | 0.00% | — | — | — |
| Tobacco use | 0 | 0.00% | — | — | — |
| Drug-resistance status | 0 | 0.00% | — | — | — |

#### 4.2 Complete-case sensitivity

We repeated the rolling landmark analysis on individuals with non-missing data for every adjustment covariate, without imputation. The complete-case set comprises 102,186 of 171,048 individuals (59.7%), including 11,528 of the 20,830 who were lost to follow-up (55.3%). At five years the complete-case standardized risk difference for all-cause mortality was 3.11 percentage points (95% CI 2.58 to 3.61), compared with 2.43 (2.07 to 2.80) pooled across the imputed datasets. The exposed-arm risk is almost unchanged, 7.17% against 7.18%; the difference arises in the comparator, 4.06% against 4.76%. Restricting to complete cases therefore raises the estimate by about a quarter, and the conclusion does not rest on the imputation model.

### 5. Calendar-period analyses

#### 5.1 Annual LTFU incidence

Annual LTFU risk rose from approximately 10% in 2013–2018 to approximately 15% in 2021–2022, coinciding with the COVID-19 pandemic (**Figure S2)**. The apparent further rise in 2023 (to approximately 22%) reflects incomplete ascertainment of the index treatment episode in the final calendar year and should be interpreted with caution.

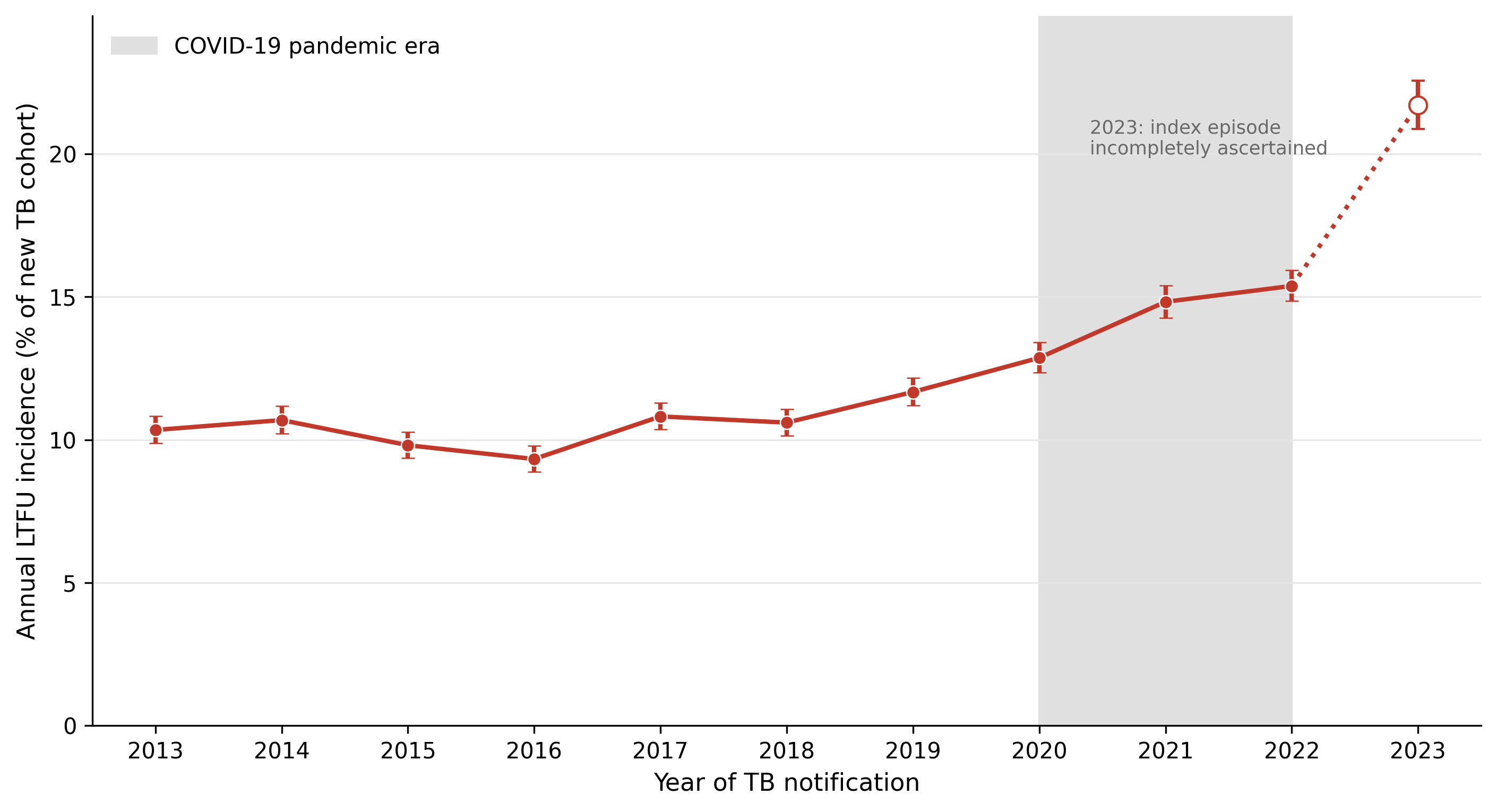

**Figure S2. Annual LTFU incidence (proportion of new TB cohort with LTFU during the index treatment episode) with 95% confidence intervals (2013–2023).** Shaded band marks the COVID-19 pandemic era (2020–2022).

### 6. Sensitivity to unmeasured confounding (E-value)

Loss to follow-up is not randomly assigned, and the covariates available in TBweb do not include markers of disease severity such as sputum smear grade, radiographic extent, CD4 count or viral load. Residual confounding is therefore a concern, and this section quantifies how strong it would have to be to account for the primary result.

We computed an E-value for the primary estimand. On the risk ratio scale, the contrast between the two assigned strategies at 60 months is 1.24 (95% CI 1.20-1.27), giving an E-value of 1.78 for the point estimate and 1.69 for the lower confidence limit. An unmeasured confounder would therefore need to be associated with both disengagement from care and mortality by a risk ratio of at least 1.78, conditional on the measured covariates, to reduce the observed association to the null, and at least 1.69 to move the confidence interval to include it.

### 7. Summary of estimands and estimates

**Table S3. Estimands, comparators and estimates. RD, risk difference; RR, risk ratio; pp, percentage points.**

| **#** | **Question being answered** | **Design** | **Time origin** | **Comparator** | **Risk, exposed** | **Risk, comparator** | **Contrast (95% CI)** | **Denominator** |
| --- | --- | --- | --- | --- | --- | --- | --- | --- |
| **1** | What would 60-month mortality be if all patients disengaged from care within the first 6 months, versus if none did? | Clone–censor–weight target trial | Treatment start (day 0) | The same patients, under the strategy of remaining in care | 11.62% | 9.40% | RD +2.23 pp (1.88 to 2.52)  RR 1.24 (1.20–1.27) | 171,048 patients, of whom 20,830 lost to follow-up |
| **2** | Among patients who disengage, what is 5-year mortality versus otherwise similar patients still in care on the same date? | Rolling landmark | Each patient's own loss-to-follow-up declaration date | Patients alive and still in care on that same date | 7.76% | 5.14% | RD +2.62 pp (2.13 to 2.91)  RR 1.51 (1.40–1.57) | 17,112 exposed |
| **3** | Does the effect depend on when in treatment disengagement occurs? | Rolling landmark, stratified by month of disengagement | Each patient's own declaration date | Patients alive and still in care on that same date | — | — | RD +0.94 to +3.27 pp across months 1 to 6; excludes zero in months 1 to 5, not distinguishable from zero in month 6 | 1,497 to 4,027 exposed per month |
| **4** | Is the excess concentrated in deaths attributed to tuberculosis? | Rolling landmark, cause-specific cumulative incidence (Aalen–Johansen) | Each patient's own declaration date | Patients alive and still in care on that same date | 2.96% / 4.80% | 1.39% / 3.75% | Tuberculosis death RD +1.57 pp (1.33 to 1.79)  Not-tuberculosis death RD +1.05 pp (0.64 to 1.30) | 17,112 exposed |
